# Assessing the impact of tungiasis on children’s quality of life in Kenya

**DOI:** 10.1101/2024.10.07.24314987

**Authors:** Lynne Elson, Berrick Otieno, Abneel K Matharu, Naomi Rithi, Esther Jebor Chongwo, Francis Mutebi, Hermann Feldmeier, Jürgen Krücken, Ulrike Fillinger, Amina Abubakar

## Abstract

Tungiasis is a neglected tropical skin disease caused by the sand flea, *Tunga penetrans* which penetrates the skin causing considerable pain and itching. In this cross-sectional study we aimed to assess its impact on the quality of life of school children in Kenya.

School pupils (198) aged 8-14 years with tungiasis were randomly selected and interviewed using a tungiasis-specific quality of life instrument (TLQI). The caregivers of each infected pupil and 199 randomly selected caregivers of uninfected pupils were interviewed using the proxy KIDscreen52^®^ to assess their child’s general health-related quality of life (HR-QoL). Generalized linear models were used to assess associations between quality-of-life variables, children’s tungiasis status and other covariables.

Among infected children, 62.4% had TLQI scores reflecting a moderate to very high impact, with no significant difference between mild and severe cases. Severe cases had a lower proxy-HR-QoL than uninfected pupils (β −21.15, 95% CI −39.63 − −2.68, p=0.025), but this was not significant in multivariable models. For the first time, this study demonstrated for children whose caregivers were depressed, tungiasis had a higher impact on their quality of life (TLQI adjusted β 0.28, 95% CI 0.08−0.49, p=0.006) and had a lower general HR-QoL (adjusted β −40.34, 95%CI −55.91− −24.76, p<0.001). Conversely, if their caregiver showed them affection, tungiasis had a lower impact on their quality of life (TLQI, adjusted β −0.45, 95% CI −0.70− −0.20, p<0.001).

Further studies are needed to investigate the interaction of tungiasis with parenting styles, the mental health of children and their caregivers and their effect on children’s well-being.

**Plain English Summary:** Tungiasis is an extremely neglected tropical skin disease which causes immense suffering among children in resource poor communities in many tropical countries. A good quality of life is considered a Right of all people and is consequently used as the most important outcome of treatment programs. The impact on quality of life by tungiasis has only been investigated in very few studies, but evidence of impact is needed to monitor interventions. Here we assessed the impact of the disease on the quality of life of school pupils using a disease-specific instrument with children and a general health-related instrument with their caregivers. The study demonstrated the considerable impact tungiasis has on the quality of life of school-age children. Tungiasis affected children’s ability to sleep and caused feelings of sadness and shame. Tungiasis had a greater impact on children whose caregivers were depressed, and they had a lower general health-related quality of life. Conversely, tungiasis had a lower impact on quality of life for children whose caregivers showed them affection. Further research is needed to investigate the interaction of tungiasis with parenting styles and the mental health of children and their caregivers and their effect on children’s well-being.

## Background

Tungiasis is a neglected tropical skin disease receiving little or no attention for research, surveillance and interventions from national governments, their partners and donors. It has only recently been added to the World Health Organization’s list of Neglected Tropical Diseases targeted for control [1].

Tungiasis is caused by female sand fleas (*Tunga penetrans)* which penetrate the skin and stay embedded for their remaining life [2]. The disease mostly affects resource-poor communities across Latin America and sub-Saharan Africa [3]. The global burden of disease is unknown, but 668 million people (304 million in East Africa) have been estimated to be at risk of infection[4]. The only national survey ever conducted, was in Kenya, and estimated the national prevalence of tungiasis to be 1.3% [5] with the regions of the country most at risk being the coastal strip, the central highlands and the west[6]. The prevalence from small surveys within affected communities has been reported to range from 7% to 60% [7–10]. Children, particularly boys[5, 10], elderly people and people with disabilities carry the highest disease burden [11].

The embedded flea causes severe morbidity with considerable pain and itching resulting from an intense inflammatory response around the rapidly growing flea [12] exacerbated by bacterial superinfection of the lesions [13]. The extent of morbidity is positively correlated with the infection intensity, and having more than 10 fleas has been classified as severe tungiasis [14]. While fleas have been found on any location of the body that has been in contact with the soil (hands, elbows, knees, buttocks) the vast majority of the embedded sand fleas are located on the feet [3].

Little is known about the further impact of tungiasis infection on a patient’s life. The pain and itching caused by inflammation have been reported to affect children’s ability to sleep [12], walk [12, 15], attend school [16, 17] and pay attention in class, resulting in poorer school exam performance [16, 17]. Recently a study in Kenya demonstrated tungiasis was associated with poor neurocognitive functioning including literacy, language, cognitive flexibility, working memory, response inhibition, numeracy and fine motor skills [18]. Two previous studies in Kenya using a tungiasis specific Quality of Life Index (TLQI) both demonstrated tungiasis significantly impacts a child’s quality of life [17, 19].

The World Health Organization (WHO) defines quality of life as “an individual’s perception of their position in the life and context of their culture and in relation to their goals, expectations, standards and concerns” [20]. Health-related quality of life reflects the impact of disease on quality of life. Due to the globally accepted right of all people to a good quality of life, clinical trials and intervention programs are now using this as an outcome measure in the treatment of various diseases such as cancer [21], diabetes [22] and lymphatic filariasis [23]. Various instruments have been developed and validated to assess quality of life in different populations, such as the WHODAS2.0 [24] and the KIDSCREEN [25] for children and adolescents, while others are designed to be specific for certain diseases, for example the DLQI for any skin disease [26] and the LFSQQ for lymphatic filariasis [27].

The objective of this study was to determine the impact of tungiasis on children’s quality of life in a large group of children in a community with a high disease burden using both a disease-specific and a general health related quality of life instrument.

## Methods

### Study design

This was a cross-sectional study of children aged 8 to 14 years, the age group most affected by tungiasis [7], plus their caregivers. Caregivers were classified as the adult who provided most of the care for the child, cooking for them, watching out for their safety and at a younger age bathing them.

### Study setting

The study reported here is part of a larger study on the ecology and impact of tungiasis in two regions of Kenya having a high prevalence of tungiasis [14]. The recruitment and data collection were conducted between February 2020 and April 2021 in the sub-counties of Msambweni and Mutuga of Kwale County on the southeast coast, and in Ugenya sub-county of Siaya County in western Kenya (Figure 1).

**Figure 1.**
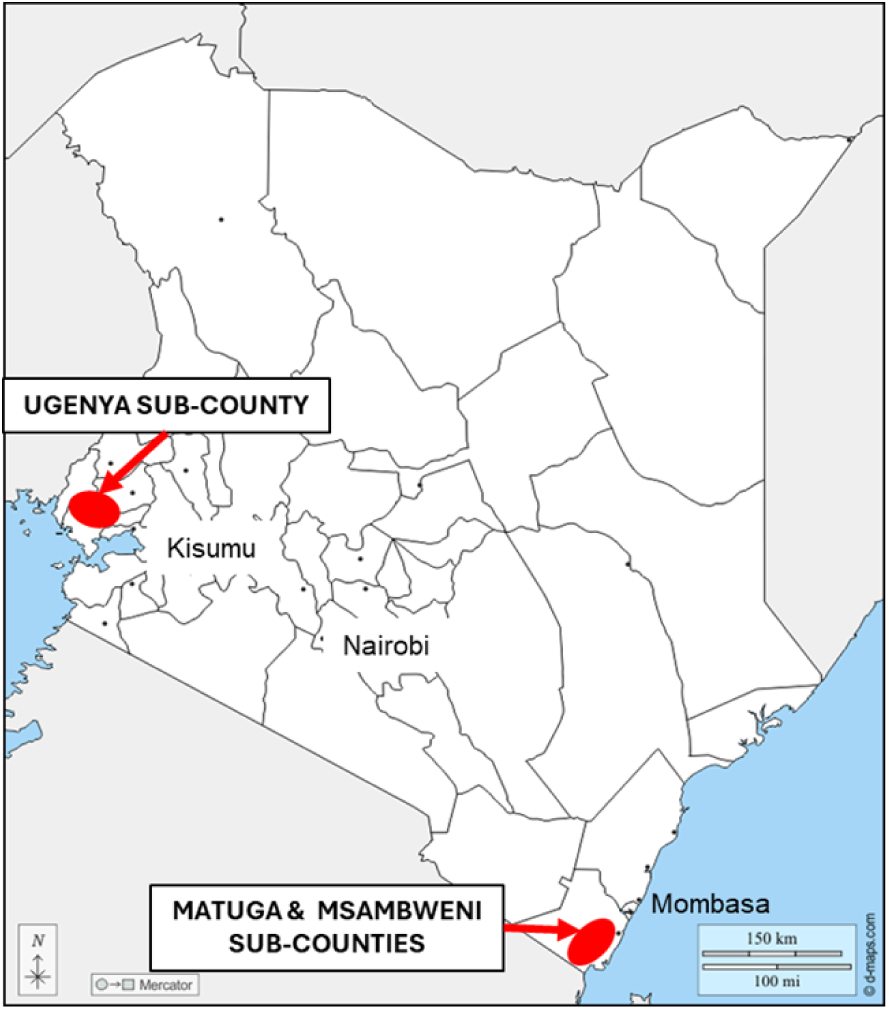
Map of Kenya to show the location of the sub-counties included in the study.

### Sample size

The same group of pupils were enrolled for several impact assessments, both quality of life reported here, and neurocognitive function which was published previously[18]. The sample size was based on the mean difference in fluency scores for infected and uninfected pupils using a previous study in a similar setting with a mean category fluency of 15.97 [10]. Assuming a common standard deviation of 2, the study aimed to enroll 506 participants (253 infected and 253 uninfected) to detect at least 0.05 difference in means at *α* = 0.05 and power of 0.8. The sample size was calculated using Stata [11]. Inclusion criteria for infected pupils were having at least one embedded flea, living in a house with an unsealed earthen floor and the availability of a caregiver to provide informed consent and to also enroll for the assessments. In addition to those described for infected pupils, uninfected pupils had to have no signs of infection and no other infected family members.

### Sampling strategy

To identify infected and uninfected pupils aged 8 to 14 years to enroll for the quality-of-life assessments, pupils in public primary schools were enrolled and examined for tungiasis. Public primary schools, 15 from Kwale and 20 from Siaya, were randomly selected from lists provided by the Departments of Education using the paper lottery method. In each school, 51 boys and 51 girls between the age of 8 and 14 years were quasi-randomly selected by lining the pupils up in three age groups; 8 and 9 years; 10 and 11 years; 12 to 14 years) and by sex within each age group. Every n^th^ (depending on the total number in the group) pupil was then selected in each sex and age group until 17 from each was reached, to a total of 102 in each school, 3,870 pupils from both counties. Each selected pupil was carefully examined for embedded fleas. For those found to be infected the number of embedded fleas (live, dead and manipulated lesions) were counted. After the examinations, using a paper-lottery method, in each school three boys and three girls were randomly selected from among tungiasis cases, and three boys and three girls from uninfected pupils, for a total of 210 cases and 210 uninfected pupils for the quality-of-life assessments. One caregiver for each of these children was also enrolled in the study.

### Procedures

The selected infected and uninfected pupils were first interviewed using structured questionnaires covering sociodemographic and psychosocial covariables about themselves, their family, their house, and school life. Only the infected pupils were then also interviewed to assess the impact of tungiasis on their quality of life using the Tungiasis Life Quality Index (TLQI). The caregivers of infected and uninfected pupils were interviewed for their perceptions of the health-related quality of life of their children using the proxy KIDSCREEN52^®^ (HR-QoL).

Through our previous work with families affected by tungiasis we had developed a hypothesis that caregiver mental health may be an underlying factor in child neglect and poor hygiene and therefore be a risk factor for tungiasis infection among children. Tungiasis has previously been associated with poor hygiene[10, 28]. Consequently, the caregivers of enrolled pupils were all assessed for their own mental health using the Patient Health Questionnaire (PHQ-9, [29]) and their stress level using the Parental Stress Scale (PSS) [30]. This also enabled us to investigate whether caregiver mental health may confound the impact of tungiasis on children’s quality of life.

### Tools for assessing the impact of tungiasis on QoL

#### The tungiasis-specific instrument (TLQI)

Life quality impairment was assessed using two different tools. One instrument was a tungiasis-specific tool inspired by the Children’s Dermatological Life Quality Index [31] which was adapted previously (TLQI) [19] and is the only quality of life instrument developed for patients with tungiasis. The instrument is provided in Supporting Information S2. The original tungiasis QLI tool had six domains each with one question, as listed in Table 1 which asked: “During the last week rate the following according to the scales (not at all, only a little, quite a lot, very much): How embarrassed or ashamed did you feel because of the jiggers, How much did the jiggers make it difficult for you to walk/run, How much do the jiggers affect your concentration in class, affect your sleep, affect your friendships, and how much are other children mean/cruel/unkind to you”.

**Table 1.** Comparison of the instrument domains.

| **TLQI-self** | **KIDSCREEN52-Proxy** |
| --- | --- |
| Mobility (walk/run) | Physical well-being |
| Feeling angry | Moods & emotions |
| Feeling sad |  |
| Shame (embarrassed) | Self-perception |
| Friendships | Social support & peers |
| Concentration due to itching | School environment |
| Bullying | Social acceptance |
| Sleep disturbance |  |
|  | Psychological well-being |
|  | Autonomy |
|  | Parent relations |
|  | Financial resources |

Based on our personal experience of tungiasis and interacting with children with tungiasis we added two new questions; “how much do the fleas make you feel sad” and “how often do you feel angry” for a total of 8 domains. The maximum total score was 24. The higher the score, the higher the impact of tungiasis and the lower the quality of life. The instrument was piloted with a few pupils in the community who did not participate in the study. This instrument is not subject to licensing and has not been systematically validated

#### Proxy-KIDSCREEN52

The second instrument assessed the caregiver’s perception of the child’s general health-related quality of life (HR-QoL) through interviews with the caregiver using a generic, standardized screening instrument for children and adolescents, the proxy-KIDSCREEN52 The instrument is provided in Supporting Information S3 [25]. This instrument has been previously validated for use in South Africa [32], Uganda and Kenya [33, 34] and is available publicly in Kiswahili [35]. This tool has 52 questions across 10 domains covering physical wellbeing, psychological wellbeing, moods and emotions, self-perception, social support and peers, social acceptance, school environment, autonomy, parent relations and financial resources. Each domain in the KIDSCREEN52 instrument has between 3 and 7 questions with each being a 5-point Likert scale.

As illustrated in Table 1, this instrument has some conceptual overlaps with the tungiasis-specific instrument. For instance, the ability to walk and run is covered by physical well-being questions while some questions under moods and emotions address feeling sad and angry. Both tools included domains assessing a child’s experience of stigma and discrimination: feeling embarrassed or ashamed are similar to questions in self-perception, affect on friendships is similar to questions in social support, and bullying is included in the questions for social acceptance. Sleep disturbance is only covered in the TLQI while psychological well-being, autonomy, financial resources and parent relations are only covered in the KIDSCREEN52 instrument.

Both tools were translated from English into the local language, Kidigo or Kiswahili in Kwale, Dholuo or Kiswahili in Siaya.

The internal consistency of each instrument was tested using Cronbach’s alpha and found both instruments had good internal consistency in this population, and all domains are valid components of the overall scores (HR-QoL 0.836, TLQI 0.866).

### Statistical analysis

All analyses were conducted in StataNow 18 BE (Stata Corp LLC, College Station, TX, USA). Disease severity was classified based on flea counts. Pupils with one to ten fleas were classified as mild, while those with more than ten fleas were classified as severe disease based on our previous study in this population [14].

#### TLQI

Initially the individual domains were assessed by calculating the number of infected pupils reporting “quite a lot” or “very much” impact (score of 2 or 3) for each domain to identify the most affected domains. Correlation of disease severity with each domain was tested using the Chi^2^ test. The total TLQI score was calculated by summing all eight domains for each pupil. For comparison with other studies, a five-level ordinal variable was generated; no impact (0-1), small (2-5), moderate (6-10), large (11-15) and very large (16-24) based on our previous studies in Kenya [17, 19] and the original children’s CDLQI [36] (see Supporting Information S4 Figure for the thresholds in a frequency histogram). Association of the TLQI score with disease severity, levels of pain and itching experienced and other possible explanatory and confounding variables were assessed using a mixed effect negative binomial model on account of the positively skewed distribution of the scores (Supporting Information S4 Figure). The school ID was included as a random effect to adjust for the clustered nature of the observations. All explanatory variables were included in a univariable analysis and those with a p-valueless than 0.2 were included in the full multivariable model. Backward elimination was used to develop the final model using Akaike information criteria (AIC). Outcomes are presented as adjusted β coefficients with 95% confidence intervals and p-values.

#### KIDSCREEN52 (HR-QoL)

The KIDSCREEN scores were processed according to the standard procedures prescribed in the manual [37]. Any negatively coded scale was inverted to be positively coded. The total score for all questions in a domain was calculated. This total domain score was then transformed into a Rasch person parameter and then to a T-value using the international norm values available online ([38], Stata code in Supporting Information S7). This transformation aims to obtain a parametric measure with a mean of 50 and a standard deviation of 10 for each domain. The domain T scores were summed up to obtain the health-related quality of life (HR-QoL) score. The lower the score the lower the quality of life. Based on the bimodal distribution of the total HR-QoL scores (Supporting Information S8), mixed effect linear models with a gaussian distribution and identity link were used to test the association of disease status and severity for each domain individually and for the T-values (HR-QoL) with pain and itching levels experienced, and independent variables. The school ID was used as a random effect to adjust for the clustered nature of the observations. Outcomes are presented as adjusted β coefficients with 95% confidence intervals and p-values.

#### Explanatory variables

The selection of possible explanatory and confounding variables to include in multivariable models was guided by a conceptual framework illustrated in Supporting Information S5. Variables associated with the disease, and/or considered to be associated with quality of life such as parent and family characteristics, parenting style, and pupil characteristics and behavior and had no conceptual overlap with the outcome variable were included (Supporting Information S6 Table). These were different for the two quality-of-life measures.

#### Missing responses

All participants were expected to provide responses to all questions (variables). Missing responses in the independent variables were ‘missing at random’ and were all less than 5% of all observations (Table S6). We therefore performed these analyses under the valid missing at random assumption, as we used likelihood approaches [39]. Final models included no less than 90% of the observations.

### Ethical considerations

This study was performed in line with the principles of the Declaration of Helsinki. The study was approved by the KEMRI Scientific and Ethics Review Committee; approval number non-KEMRI 644. Permission to conduct the study was also obtained from the county and sub-county Health Management Teams and the Department of Education. Informed consent was obtained from the school Head Teachers and PTA chairperson. Caregivers of the pupils selected for interviews gave informed consent for their own involvement as well as for their child, all ages enrolled. In addition, pupils aged 12 to 14 years themselves gave signed informed assent.

All data were collected on PIN protected electronic tablets, stored on password protected databases on the servers of the International Centre of Insect Physiology and Ecology, Nairobi. Personal identifiers were removed from analyses data sets.

All pupils with tungiasis were treated by the community health workers or the local health facility using benzyl benzoate, which was the product chosen by the county health managers and provided by the study free of charge.

## Results

### Pupil participants

A total of 3,870 pupils were examined for tungiasis and 481 were found to be infected as described previously [14] and illustrated in the flow chart in Figure 2. From these, 210 infected and 210 uninfected pupils were stratified randomly selected to participate, but only 198 infected and 199 uninfected pupils and their caregivers consented to enroll into the quality-of-life study and were included in analyses (Figure 2). As intended by the selection criteria, they were evenly distributed between Kwale (195) and Siaya (202), and between boys (227) and girls (168). Of the infected pupils, 103 (52.0%) had severe disease (more than 10 fleas). Distribution of participants by all covariables included in the multivariable models are detailed in the Supporting Information S6.

**Figure 2.**
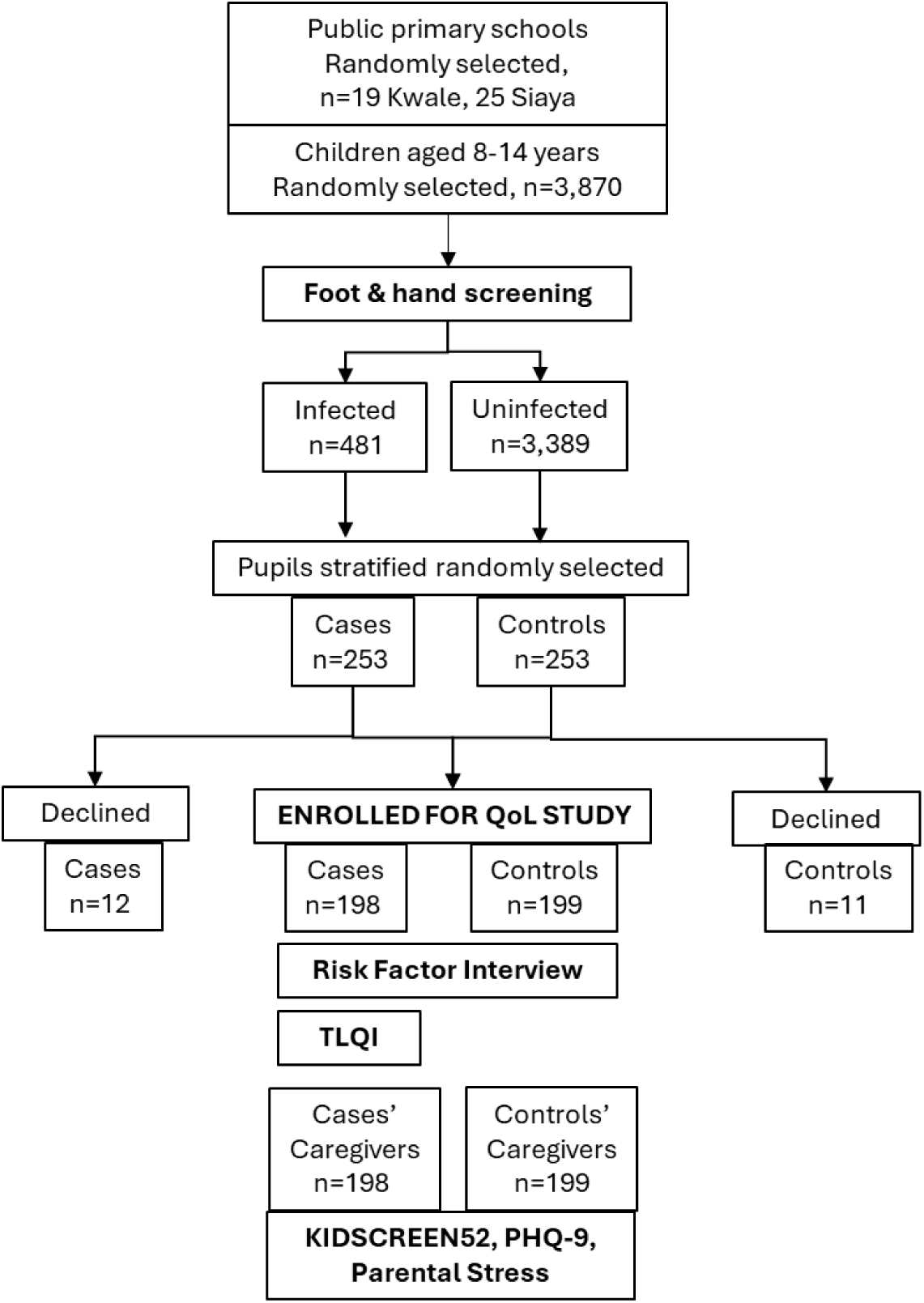
Flow chart of pupils examined for tungiasis, selected, and enrolled for the quality-of-life study.

### The two instruments

The TLQI was only conducted with 198 infected children. The Proxy-KIDSCREEN52 was conducted with the caregivers of the 198 infected children, plus caregivers of the 199 uninfected children.

### The tungiasis life quality index

With this instrument we assessed the level of impact of tungiasis on the disease specific quality of life of infected pupils and compare pupils with mild disease to those with severe disease. The higher the score the greater the impact and the lower the quality of life.

The percent of pupils who reported experiencing “quite a lot” or “very much” impact was highest for disturbed sleep, feelings of shame and sadness while those domains with the lowest percent of pupils experiencing these levels of impact were friendships and bullying (Table 2).

**Table 2.** Chi^2^ correlation of TLQI domains with disease severity. Percent of pupils who reported experiencing “quite a lot” or “very much” for each domain by disease severity.

| Domain | All cases<br>n=196 (%) | Mild<br>n=94 (%) | Severe<br>n=102 (%) | p-value |
| --- | --- | --- | --- | --- |
| Sleep disturbance | 38.3 | 30.9 | 45.1 | 0.040 |
| Feeling ashamed | 35.7 | 28.7 | 42.2 | 0.050 |
| Feeling sad | 33.7 | 24.5 | 42.4 | 0.009 |
| Concentration in class | 29.1 | 26.6 | 31.4 | 0.462 |
| Mobility | 27 | 21.3 | 32.4 | 0.081 |
| Feeling angry | 26.5 | 19.2 | 33.3 | 0.025 |
| Bullying | 23 | 21.3 | 24.5 | 0.591 |
| Friendships | 19.9 | 11.7 | 27.5 | 0.006 |

A higher percentage of pupils with severe tungiasis reported higher levels of impact than those with mild disease in all domains (p<0.050) except concentration in school and bullying.

When the scores of all domains were combined for each child, the median TLQI score for all pupils was 7 (interquartile range (IQR) 4−12.5). The median score for mild cases was 7 (IQR 3−12) and for severe cases was 8 (IQR 5−13). When grouped into five categories of impact, of all cases 62.4% reported experiencing a moderate to very large impact on their quality of life. Of the mild cases, 54.7% experienced this level of impact and 68.4% of severe cases (Figure 3). Among the severe cases 31.6% reported no or only a small impact and 20% of mild cases reported a very large impact.

**Figure 3.**
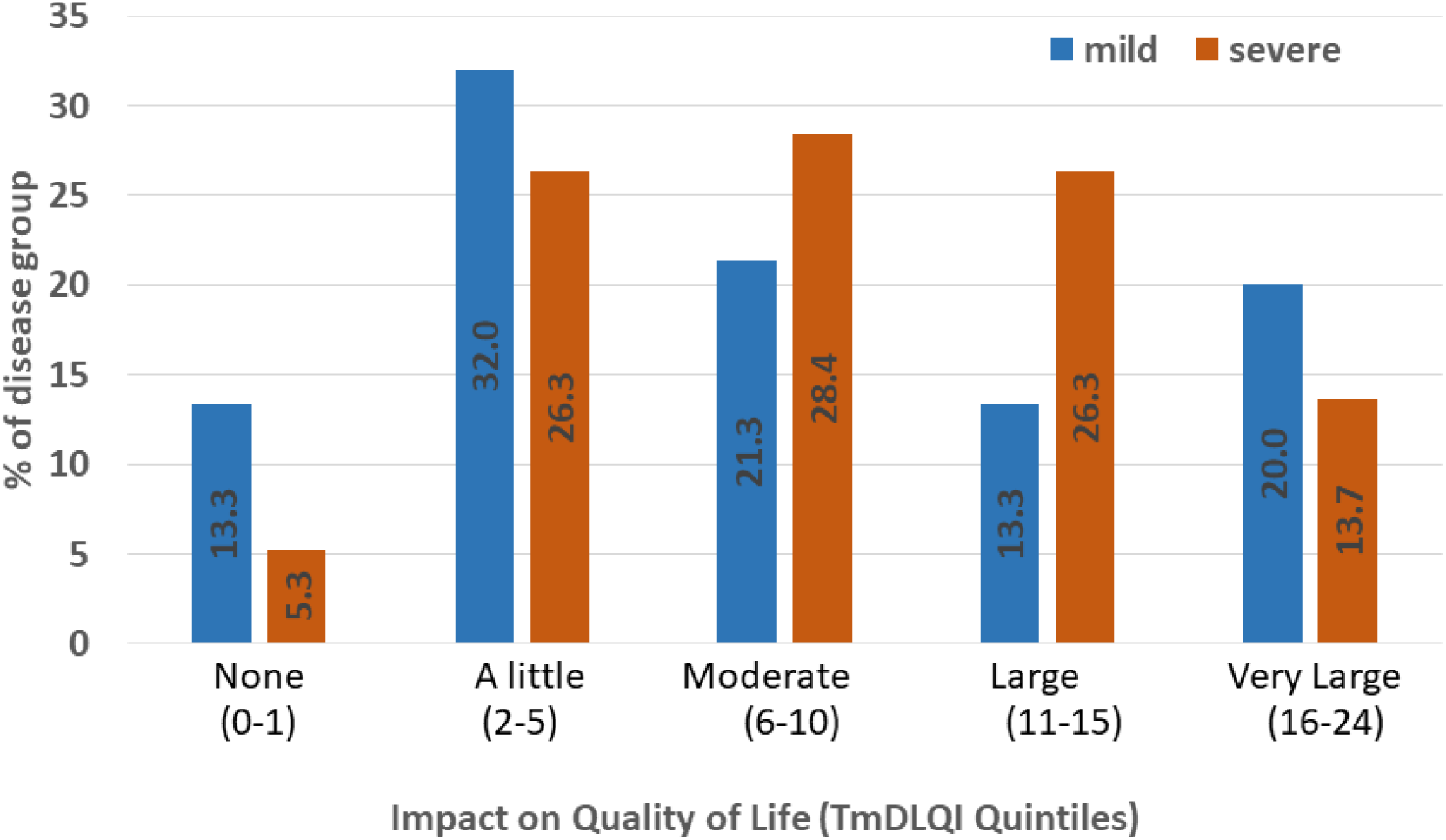
Frequency distribution of pupils across the five TLQI categories reflecting impact on quality of life by disease severity. Blue bars: pupils with mild disease, orange bars: pupils with severe disease.

In the univariable regression analyses for TLQI we found that severe tungiasis was associated with a higher TLQI (β 0.24, 95% CI 0.03−0.45, p=0.027; Supporting Information S9). Once adjusted for other covariables in the final multivariable model this association remained (adjusted β 0.27, 95% CI 0.07−0.48, p=0.009, Table 3).

**Table 3.** Mixed effect negative binomial regression analysis of associations of TLQI with tungiasis severity and covariables. School ID used as random effect.

| Variables | Categories | N<sup>1</sup> | Full model (n=187) |  | Final model (n=189) |  |
| --- | --- | --- | --- | --- | --- | --- |
| | | | Adjusted $\beta$ Coefficient<br>(95% CI)<sup>2</sup> | P<sup>3</sup> | Adjusted $\beta$ Coefficient<br>(95% CI) | P |
| Tungiasis severity | Mild | 93 | ref |  | ref |  |
|  | Severe | 102 | 0.24 (0.03–0.44) | 0.027 | 0.27 (0.07–0.48) | 0.009 |
| Region | Kwale | 95 | ref |  |  |  |
|  | Siaya | 100 | -0.07 (-0.34–0.20) | 0.634 |  |  |
| Sex | Female | 56 | ref |  |  |  |
|  | Male | 139 | 0.04 (-0.18–0.26) | 0.720 |  |  |
| Pupil age |  | 196 | 0.02 (-0.04–0.08) | 0.536 |  |  |
| Adults live with | Both parents | 121 | ref |  |  |  |
|  | Others | 77 | -0.03 (-0.39–0.32) | 0.861 |  |  |
| Primary caregiver | Mother | 145 | ref |  | ref |  |
|  | Other adult | 52 | 0.37 (-0.03–0.76) | 0.067 | 0.38 (0.06–0.71) | 0.021 |
| Mother's education | Secondary | 68 | ref |  |  |  |
|  | None | 16 | 0.16 (-0.22–0.53) | 0.415 |  |  |
|  | Primary | 87 | 0.00 (-0.29–0.29) | 0.985 |  |  |
|  | Don't know | 27 | 0.22 (-0.15–0.59) | 0.250 |  |  |
| Father's education | Secondary | 62 | ref |  |  |  |
|  | None | 11 | -0.68 (-1.19– -0.18) | 0.008 |  |  |
|  | Primary | 88 | -0.11 (-0.40–0.17) | 0.434 |  |  |
|  | Don't know | 36 | -0.29 (-0.62–0.04) | 0.089 |  |  |
| Mother away a lot | No | 130 | ref |  | ref |  |
|  | Yes | 63 | -0.37 (-0.70– -0.04) | 0.030 | -0.45 (-0.77– -0.14) | 0.005 |
| Parents attend school meetings | Often | 83 | ref |  | ref |  |
|  | Not often | 114 | -0.27 (-0.49– -0.06) | 0.014 | -0.30 (-0.50– -0.11) | 0.003 |
| Parents check homework | Never | 35 | ref |  |  |  |
|  | Sometimes | 163 | -0.08 (-0.36–0.20) | 0.582 |  |  |
| Family member ill some months | No | 117 | ref |  |  |  |
|  | Yes | 78 | 0.23 (-0.01–0.47) | 0.059 |  |  |
| Caregiver depressed | No | 114 | ref |  | ref |  |
|  | Yes | 81 | 0.31 (0.10–0.53) | 0.005 | 0.28 (0.08–0.49) | 0.006 |
| Caregiver time spend with child | A little | 146 | ref |  |  |  |
|  | A lot | 47 | -0.10 (-0.35–0.16) | 0.466 |  |  |
| Child fears a family member | No | 130 | ref |  |  |  |
|  | Yes | 65 | 0.01 (-0.24–0.26) | 0.913 |  |  |
| Caregiver hugs child | No | 139 | ref |  | ref |  |
|  | Yes | 55 | -0.37 (-0.64– -0.10) | 0.007 | -0.45 (-0.70– -0.20) | <0.001 |
| Discipline style | Other | 62 | ref |  |  |  |
|  | Beaten | 136 | 0.03 (-0.20–0.26) | 0.788 |  |  |
| HHH<sup>4</sup> sex | Female | 42 | ref |  |  |  |
|  | Male | 151 | 0.26 (-0.09–0.61) | 0.143 |  |  |
| HHH age |  | 196 | 0.00 (-0.01–0.01) | 0.638 |  |  |
| Caregiver sex | Female | 169 | ref |  |  |  |
|  | Male | 24 | 0.00 (-0.37–0.37) | 0.990 |  |  |
| Caregiver age |  | 196 | 0.00 (-0.01–0.02) | 0.648 |  |  |

<sup>1</sup>number, <sup>2</sup>confidence interval, <sup>3</sup>p-value, <sup>4</sup>head of household

In the final multivariable model other covariables were also associated with the TLQI. Children whose caregivers were classified as depressed had a higher TLQI (adjusted β 0.28, 95% CI 0.08−0.49, p=0.006) than those whose caregiver was not depressed, which means they experienced a higher impact from tungiasis than those whose caregivers were not depressed (Table 3). Other factors associated with a higher TLQI included if a child’s main caregiver was not their mother (adjusted β 0.38, 95% CI 0.06−0.71, p=0.021). Conversely, children whose caregiver reported they hug and cuddle their child had a lower TLQI than children whose caregiver did not, which means they experienced a lower impact of tungiasis (adjusted β −0.45, 95% CI −0.70− −0.20, p<0.001). Additionally, parents who only attended school meetings sometimes vs. often (adjusted β −0.30, 95% CI −0.50− −0.11, p=0.003), and if a child’s mother was away a lot (adjusted β −0.45, 95% CI −0.77− −0.14, p=0.005) were associated with lower TLQI scores (Table 3).

### General health related quality of life assessed by the caregiver (KIDscreen52®)

With this instrument we compared the scores of uninfected pupils with pupils having mild and severe tungiasis separately. The lower the score the lower the health-related quality of life.

The domains with lowest mean scores for all pupil groups were financial resources, social acceptance, and physical well-being (Table 4). The financial resources were extremely low irrespective of disease status. Compared to uninfected children, those with severe tungiasis were associated with lower scores in the domains of: psychological well-being (β - 3.53, 95% CI −6.74 − −0.32, p=0.031), moods and emotions (β −4.16, 95% CI −7.05 − −1.27, p=0.005), self-perception (β −4.35, 95% CI −6.99 − −1.71, p=0.001), school environment (β - 3.05, 95% CI −5.88 − −0.22, p=0.034) and social acceptance (β-5.86, 95% CI-9.51 − −2.21, p=0.002) (Table 4).

**Table 4.** Univariable generalized linear regression analysis for the individual domains of HR-QoL (KIDSCREEN52) by tungiasis severity.

| | Disease group | Mean | SD<sup>1</sup> | $\beta$<br>Coefficient | 95% CI<sup>2</sup> | | P |
| --- | --- | --- | --- | --- | --- | --- | --- |
| Physical well-being | Uninfected | 51.9 | 12.6 |  |  |  |  |
|  | Mild | 53.8 | 13.7 | 1.90 | -1.27 | 5.07 | 0.240 |
|  | Severe | 49.0 | 13.2 | -2.83 | -5.91 | 0.26 | 0.073 |
| Psychological well-being | Uninfected | 52.0 | 13.0 |  |  |  |  |
|  | Mild | 53.6 | 14.1 | 1.67 | -1.63 | 4.97 | 0.321 |
|  | Severe | 48.4 | 14.1 | -3.53 | -6.74 | -0.32 | 0.031 |
| Moods & emotions | Uninfected | 59.9 | 11.2 |  |  |  |  |
|  | Mild | 58.3 | 13.4 | -1.65 | -4.62 | 1.31 | 0.275 |
|  | Severe | 55.8 | 12.9 | -4.16 | -7.05 | -1.27 | 0.005 |
| Self-perception | Uninfected | 54.7 | 10.9 |  |  |  |  |
|  | Mild | 55.0 | 11.4 | 0.34 | -2.37 | 3.05 | 0.806 |
|  | Severe | 50.3 | 11.3 | -4.35 | -6.99 | -1.71 | 0.001 |
| Autonomy | Uninfected | 48.3 | 13.4 |  |  |  |  |
|  | Mild | 48.9 | 14.1 | 0.60 | -2.65 | 3.84 | 0.719 |
|  | Severe | 48.5 | 12.4 | 0.22 | -2.94 | 3.38 | 0.892 |
| Parent relations | Uninfected | 53.8 | 12.0 |  |  |  |  |
|  | Mild | 55.8 | 12.5 | 2.00 | -1.00 | 5.00 | 0.191 |
|  | Severe | 52.6 | 12.7 | -1.20 | -4.12 | 1.72 | 0.422 |
| Financial Resources | Uninfected | 28.1 | 7.3 |  |  |  |  |
|  | Mild | 27.8 | 8.9 | -0.31 | -2.24 | 1.61 | 0.75 |
|  | Severe | 27.6 | 8.0 | -0.48 | -2.35 | 1.40 | 0.618 |
| Social support & peers | Uninfected | 46.7 | 14.0 |  |  |  |  |
|  | Mild | 50.6 | 15.2 | 3.99 | 0.52 | 7.45 | 0.024 |
|  | Severe | 48.9 | 13.8 | 2.20 | -1.17 | 5.57 | 0.201 |
| School Environment | Uninfected | 58.4 | 11.5 |  |  |  |  |
|  | Mild | 60.3 | 12.1 | 1.91 | -1.00 | 4.81 | 0.198 |
|  | Severe | 55.3 | 12.6 | -3.05 | -5.88 | -0.22 | 0.034 |
| Social Acceptance | Uninfected | 42.7 | 13.9 |  |  |  |  |
|  | Mild | 41.4 | 17.5 | -1.26 | -5.01 | 2.49 | 0.510 |
|  | Severe | 36.8 | 16.1 | -5.86 | -9.51 | -2.21 | 0.002 |

<sup>1</sup> standard deviation, <sup>2</sup> confidence interval

When all the domain scores were combined into the overall HR-QoL score for each child, the mean scores were lowest for children with severe tungiasis 473.4 (standard deviation (sd 82.0) compared to 496.5 (sd 77.8) for uninfected children (Table 5). In univariable analysis there was no association with infection when mild and severe cases were combined (β −7.93, 95% CI −23.16 − 7.29, p=0.307), but when cases were separated by disease severity, severe disease was associated with a lower HR-QoL relative to no infection (β −21.15, 95% CI −39.63 − −2.68, p=0.025, Supporting Information Table S10) There was no association of the HR-QoL with mild tungiasis relative to uninfected children. In the multivariable model adjusting for covariables, the association between HR-QoL and severe tungiasis was no longer statistically significant (adjusted β −13.28, 95% CI −30.71−4.15, p=0.135, Table 5).

**Table 5.**
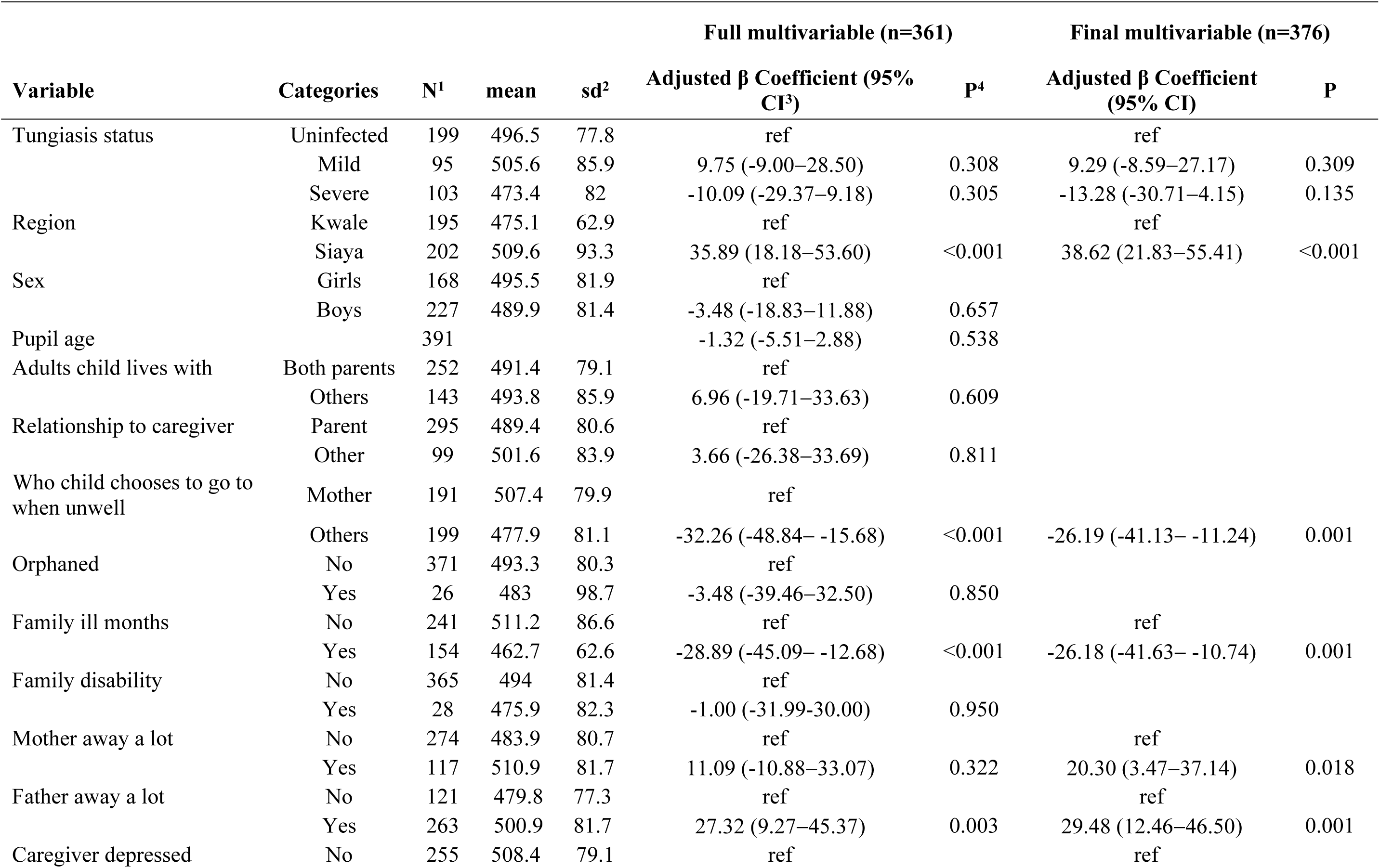
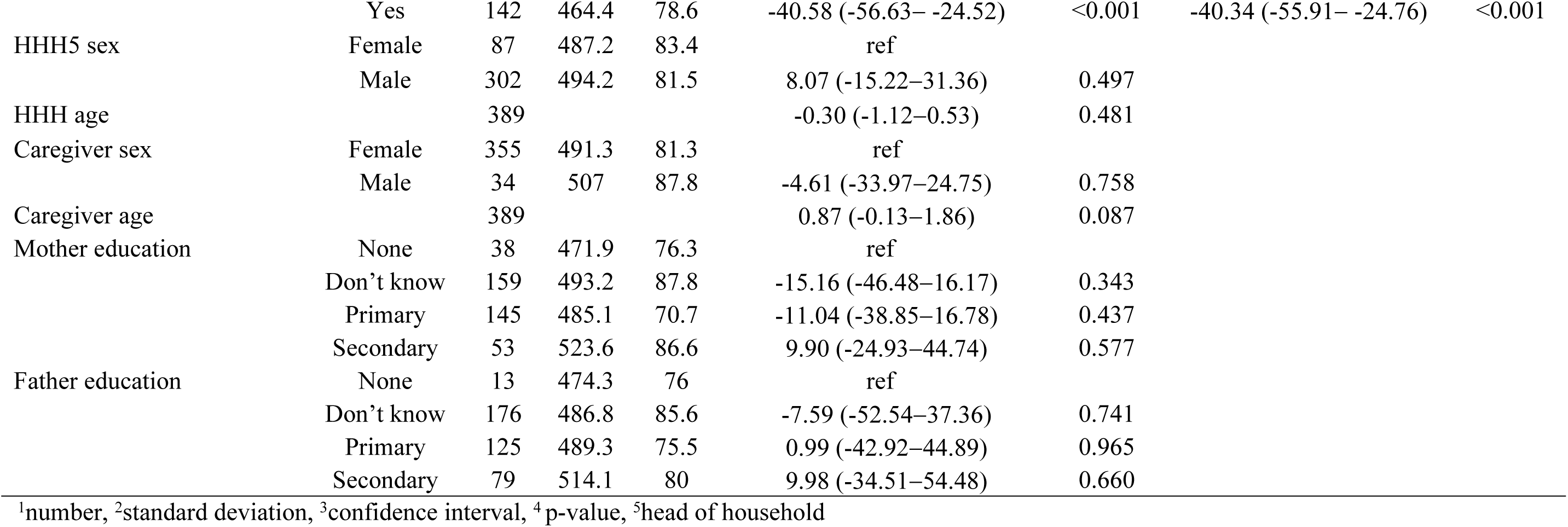
Mixed effect linear regression analysis of associations of disease status and covariables with HR-QoL (KIDSCREEN52) using school.

Variables that were associated with a higher HR-QoL included: residing in Siaya county (adjusted β 38.62, 95% CI 21.83−55.41, p<001); if the father was away a lot (adjusted β 29.48, 95% CI 12.46 − 46.50, p=0.001) and if the mother was away a lot (adjusted β 20.30, 95% CI 3.47 − 37.14, p=0.018) (Table 5). Variables associated with a lower HR-QoL included: if the caregiver was classified as depressed (adjusted β −40.34, 95%CI −55.91-−24.76, p<0.001), if a family member was chronically ill (adjusted β - 26.18, 95% CI - 41.63− −10.74, p<0.001,) and if the child chooses to go to an adult other than their parent when feeling unwell (adjusted β 26.19, 95% CI −41.13− −11.24, p=0.001)(Table 5).

### Pain and itching

Two of the major symptoms of tungiasis are pain and itching caused by the inflammation induced by embedded fleas[12]. To explore whether pain and itching levels might be associated with the quality-of-life measures, each infected child was asked about their experience of pain and itching in their feet as part of the interview for the TLQI. In univariable regression analysis the higher the category of pain the higher the TLQI (β 0.36, 95% CI 0.27−0.45, p=0.001) and likewise for itching (β 0.35, 95% CI 0.26−0.45, p=0.001). There was no significant association of pain and itching with the HR-QoL assessment conducted by their caregiver (Table 6).

**Table 6.** Association of pain and itching with quality-of-life scores for all infected children (n=198)

| TLQI<sup>1</sup> | $\beta$ Coefficient<sup>2</sup> | 95% CI<sup>3</sup> | | P |
| --- | --- | --- | --- | --- |
| pain | 0.36 | 0.27 | 0.45 | <0.001 |
| itch | 0.35 | 0.26 | 0.45 | <0.001 |
| **HR-QoL<sup>4</sup>** |  |  |  |  |
| pain | -6.59 | -18.01 | 4.83 | 0.258 |
| itch | -8.11 | -19.62 | 3.40 | 0.167 |

<sup>1</sup>Tungiasis quality of life index.
<sup>2</sup> $\beta$ coefficient from mixed effect linear regression model with school ID as random effect
<sup>3</sup>confidence interval
<sup>4</sup>Health related quality of life score (KIDSCREEN52)

## Discussion

In this study we set out to determine whether tungiasis had an impact on children’s quality of life and whether there was any difference between mild and severe disease, using a disease-specific, and a proxy general-health related instrument. This study reinforced previous findings that tungiasis, particularly severe disease, impacts children’s quality of life considerably and identified that children feel the most impact on their sleep and makes them feel ashamed and sad. The most important new finding is that caregiver mental health was strongly associated with both measures (tungiasis-specific and general health) of quality of life. Children whose caregivers were classified as depressed had a higher TLQI and lower proxy HR-QoL. Caregiver depression has previously been associated with poor physical and cognitive development and life satisfaction among children[40, 41]. A depressed caregiver gives less attention to their children[42], including their children’s personal hygiene [43]. Low frequency of bathing and soap-use by children have been shown to be risk factors for tungiasis [7, 10]. Not only could caregiver depression put children at greater risk for infections such as tungiasis (not assessed here), but also to being more vulnerable to the impact of disease on their quality of life, as seen here.

An important component of parenting is the display of affection through physical contact and hugging the child for their psychological well-being and development, absence of which can have a lifelong impact on the child [44]. In the present study, if a caregiver reported that they currently hug the child, their child had a lower TLQI. Another variable reflecting parent attention and care for the child, “attending school meetings”, was also negatively associated with the TLQI, that is, more parent attention in a child’s life was associated with tungiasis having a lower impact on the child. These findings suggest that a caregiver can reduce the impact tungiasis has on their child by changing their parenting style even if they cannot eliminate the disease. This should be investigated further using validated instruments for parenting styles along with developing an intervention incorporating parenting styles

The proxy HR-QoL did not include a sleep disturbance domain, but this was the most affected domain in the TLQI. Sleep deprivation is a common symptom of parasitic skin diseases; tungiasis [17, 19], cutaneous larva migrans [45] and scabies [46] which is concerning since it impacts cognitive performance [47], learning and memory formation [48], and can lead to depression, other psychological disorders and behavioral changes [49]. In fact, assessments done with these same children in this study showed that tungiasis was associated with significant cognitive impacts [18]. Further studies with a larger sample size are needed to confirm the associations of tungiasis with poor maternal and child mental health and cognitive development and to explore possible causal pathways.

There was considerable discrepancy in the responses to the social acceptance (bullying) domain in the two instruments. It was the most affected domain according to caregivers, while it was the least affected domain according to the children themselves. Past studies have found it is quite common for there to be low correlation between the child and caregiver assessments [50, 51]. Children themselves may not want to admit to a stranger that they are being bullied and excluded, while their caregivers are aware of the situation and did not have the same inhibitions. Alternatively, the caregiver may think that the child is being bullied when in fact they were not. Poor correlation between caregiver and child assessments may also be due to poor parent-child relationships; the caregiver not having much knowledge of the child; circumstances and denial on the part of either party; and the pressure on each to give socially acceptable responses and the caregiver’s stress levels [52]. The issue of bullying needs to be investigated further.

The median TLQI score (7, IQR 4−12.5) for all patients and the proportion of children who reported a moderate to very high impact (62.4%) in the current study was similar to that seen in the recent nine-county survey in Kenya (9, IQR 4−13 and 68.5%) [17]. However, these were considerably lower than the previous study in Kilifi County, Kenya (78%) [19], and those with cutaneous larva migrans in Brazil (71%) [45]. The lower levels of impact may be purely due to the tools having a different number of domains (eight and six respectively) and grouping the scores differently for the quintiles. However, this difference could also be due to cultural differences between the study populations. This suggests that there is need for a standardized instrument and analysis processes to be used for tungiasis studies and perhaps all parasitic skin diseases. The good internal consistency and frequency of a higher rating among children with severe disease, suggests the addition of the two extra variables, feelings of anger and sadness were probably appropriate, but the whole instrument requires systematic validation.

What is unexpected in this study is that 31.6% of children with severe disease reported little or no impact and 20% of children with mild disease reported high levels of impact. Kehr et al [53] suggest the high level of keratinization of the feet associated with fleas in chronic disease means some children may experience less impact. Another possible explanation is that children’s responses are influenced by their past experience, familial, cultural and cognitive factors that have been shown to play a role in pain perception and expression [54].

The TLQI was positively associated with levels of pain and itching which is perhaps not unexpected since higher levels of pain are likely to make it more difficult to walk and sleep. The question relating to concentration in class is specifically asked in relation to the itching caused by the fleas, and itching in other chronic skin diseases has been shown to worsen at night, causing sleep disturbance [55, 56]. The lack of association of the children’s pain and itching levels with the HR-QoL is perhaps a reflection of the fact that it was a proxy assessment of the child’s quality of life and that pain and itching are not associated with its various domains when assessed by the caregiver rather than the child. This result combined with the finding that severe disease was not associated with the HR-QoL suggests it is not a suitable instrument to use to assess the impact of tungiasis in a child’s quality of life.

### Study limitations

One possible limitation of the study was that while there was some conceptual overlap, the two instruments chosen for the study were quite different from each other, one being disease-specific, implemented with affected children while the other addressed general health and was implemented with the caregiver. Ideally, we would have included the KIDSCREEN52 self-assessment instrument for the child’s general HR-QoL. However, we did not wish to overburden the child participants who were also completing extensive cognitive assessments at the same visit [18], so the proxy instrument was used instead. The good internal consistency (Cronbach’s alpha >0.8) in both instruments suggests they accurately measured the tungiasis- or health-related quality of life respectively, in this study population. The good internal consistency of the HR-QoL also indicates the international norm values for the Rasch person parameters used for the KIDSCREEN52 transformations were suitable for this population. At the domain level both instruments found children with severe tungiasis experienced more sadness/depressive moods and shame/low self-esteem than children with mild tungiasis (less than 10 fleas).

Another limitation related to the instruments was that the wording of the questions in the TLQI was ambiguous, particularly that relating to friendships. It is not clear whether the responses from the children indicate a positive or negative impact on their friendships. In this study the instrument appears to be suitable, but it could still be improved and needs systematic validation.

In conclusion, the cross-sectional study design meant that we have been able to demonstrate an association of severe tungiasis with low disease-specific quality of life and of other variables with both measures (TLQI and HR-QoL), but further studies are needed to investigate the causality of these associations. Since there was no association of tungiasis severity with the generalized HR-QoL as assessed by the proxy-KIDSCREEN52 instrument suggests this may not be an appropriate instrument to assess the impact of tungiasis on quality of life. The role of caregiver mental health and parenting style, as well as children’s behaviors in both disease incidence and quality of life of children in these resource poor communities needs to be further investigated with the aim to develop holistic intervention programs aiming to improve quality of life of both caregivers and children.

## Data Availability

The datasets supporting the conclusions of this article are available on the KEMRI-Wellcome Trust Research Programme (KWTRP) Research Data Repository at Harvard Dataverse through the following link: https://doi.org/10.7910/DVN/TKXWRF

## Supporting information

S1. Table. STROBE checklist.

S2. The TLQI questionnaire.

S3. KIDSCREEN52 parental assessment of child quality of life questionnaire.

S4. Figure. Frequency histogram of the TLQI scores. Red lines indicate the thresholds used to create the five-level ordinal variable.

S5. Table. Participant Distribution by dependent and independent variables and disease status.

S6. Figure. Conceptual Framework for the association of tungiasis with Quality of Life

S7. Stata code for transforming KIDSCREEN52 scores.

S8. Figure. Frequency histogram of the HR-QoL T-values.

S9. Table. Bivariable linear regression analysis for HR-QoL

## Statements and declarations

### Author contributions

Conceptualization, LE, AA, UF, FM, HF & JK; Methodology, LE, AA, EC, FM, UF & JK; Formal Analysis, LE, BO, UF, JK, AA; Investigation, AM, BO, NR, EC; Resources, UF, AA, JK.; Data Curation, LE, AM, NR, BO.; Writing – Original Draft Preparation, LE.; Writing – Review & Editing, all; Visualization, LE.; Supervision, AA, UF, JK; Project Administration, UF, JK; Funding Acquisition, LE, AA, HF, UF, JK.

## Acknowledgements

We are grateful to the caregivers and pupils who participated, the school Parent Teacher Associations and Head Teachers who allowed us to work in their schools, the sub-county and county Directors of Health and Education who gave their approval for the study. We are grateful to Ibrahim Kiche for logistics support, Andrew Espira for the set up and maintenance of the RedCap data collection tools and database and Paul Mwangi for assistance with transforming and interpreting the KIDSCREEN52 data. We thank the field enumerators for their long hard days of work.

## Ethics approval and consent to participate

The study was approved by the KEMRI Scientific and Ethics Review Committee (approval number non-KEMRI 644) as well as the Ethikkommission of the Charité Berlin (reference number EA2/100/16). During the community entry phase, a presentation was made to the county and sub-county health management teams and the department of education in both counties to obtain their approval. In each school a meeting was held with the school parent teachers’ association (PTA) or management board to obtain their permission to conduct the survey in their school. The head teacher and PTA chairperson signed the consent form on behalf of the parents and school. Each child gave verbal assent. Community health workers were hired and trained in each school to assist and be the link with the community emphasizing that participation was completely voluntary, and subjects had the opportunity to withdraw from the study at any point. Before the interviews, informed assent and consent were obtained from the children and their caregivers, respectively. All assessments and interviews were conducted in empty classrooms for privacy.

All data were collected on PIN protected electronic tablets, stored on password protected RedCap databases on the icipe servers. Data were analyzed after export to Excel spreadsheets without inclusion of personal identifiers.

All pupils with tungiasis were referred for treatment to the community health workers or the local health facility using benzyl benzoate, chosen by the county health managers, and provided by the study. For those with secondary bacterial infection and other illnesses requiring treatment, a referral was made to the nearest health facility.

## Consent for publication

Not applicable

